# The effect of smoking on multiple sclerosis: a mendelian randomization study

**DOI:** 10.1101/2020.06.24.20138834

**Authors:** Ruth E Mitchell, Kirsty Bates, Robyn E Wootton, Adil Harroud, J. Brent Richards, George Davey Smith, Marcus R Munafò

## Abstract

The causes of multiple sclerosis (MS) remain unknown. Smoking has been associated with MS in observational studies and is often thought of as an environmental risk factor. We used two-sample Mendelian Randomization (MR) to examined whether this association is causal using genetic variants identified in genome-wide association studies (GWAS) as associated with smoking. We assessed both smoking initiation and lifetime smoking behaviour (which captures smoking duration, heaviness and cessation). There was very limited evidence for a meaningful effect of smoking on MS susceptibility was measured using summary statistics from the International Multiple Sclerosis Genetics Consortium (IMSGC) meta-analysis, including 14,802 cases and 26,703 controls. There was no clear evidence for an effect of smoking on the risk of developing MS (smoking initiation: odds ratio [OR] 1.03, 95% confidence interval [CI] 0.92-1.61; lifetime smoking: OR 1.10, 95% CI 0.87-1.40). These findings suggest that smoking does not have a detrimental consequence on MS susceptibility. Further work is needed to determine the causal effect of smoking on MS progression.

## Background

Smoking is an avoidable environmental cause to many life-threatening diseases such as lung cancer, heart and respiratory disorders (1,2). There is emerging evidence linking cigarette smoke to conditions negatively affecting the central nervous system (CNS), like multiple sclerosis (MS) (3,4). MS is a chronic neurological disorder causing autoimmune breakdown of the myelin sheath surrounding axons in the CNS (5). The disease is characterised by periods of disease activity followed by remission and/or progressive neurological decline, resulting in increasing disability (6). Like most autoimmune conditions, there is no known specific cause; however, we know there is an interaction between genetic and environmental factors in susceptible individuals, that go on to develop the disorder (7). Unfortunately, there is no cure for MS(8), and people diagnosed often live with extreme disability (9). There are emerging treatments aimed at modifying the disease course (10), but they are not universally effective particularly with regards to the progressive form of the disease. Therefore, it is important to continue targeting prevention by means of establishing causal links. Evidence from observational epidemiological studies suggests that smoking increases MS risk (11). It is hypothesised from experimental studies that exposure to chemicals in cigarette smoke alter the immune cell balance in the lung (12,13) which in turn can lead to generalised pro-inflammatory effects that trigger autoimmunity (14,15), in genetically susceptible individuals (16,17). In addition, cigarette chemicals contribute mechanistically to MS pathobiology. Specifically, nicotine increases the permeability of the blood-brain barrier (18); cyanide contributes to demyelination (19); and nitric oxide causes degeneration of axons (20). There is evidence for an association between smoking and worsening symptoms, number of relapses, lesion load on MRI, brain atrophy rate (15) and the rapidity of disability progression in MS patients (4,21,22).

However, it is hard to make causal inferences from observational studies which can be biased by issues of reverse causation and residual confounding. One method which can be used to reduce these sources of bias is Mendelian Randomization (MR) (23). MR can be implemented through instrumental variable analysis that uses genetic variants to proxy the exposure (e.g, smoking) and estimate a causal effect of that exposure on the outcome (e.g, MS). The MR method makes three important assumptions: 1) the genetic variants must robustly predict the exposure, 2) the genetic variants must not be associated with any confounders and 3) the genetic variants must only affect the outcome through the exposure (24). To satisfy the first assumption we selected the most recently available genetic instruments from previously conducted genome-wide association studies (GWAS) associated with smoking behaviour (smoking initiation (25) and lifetime smoking (26)) that can be implemented in a two-sample MR context (Fig 1). The latter two assumptions can be violated by horizontal pleiotropy which occurs when the genetic variants affect the outcome other than through the exposure. We test for this possibility using multiple sensitivity analyses.

**Fig 1.**
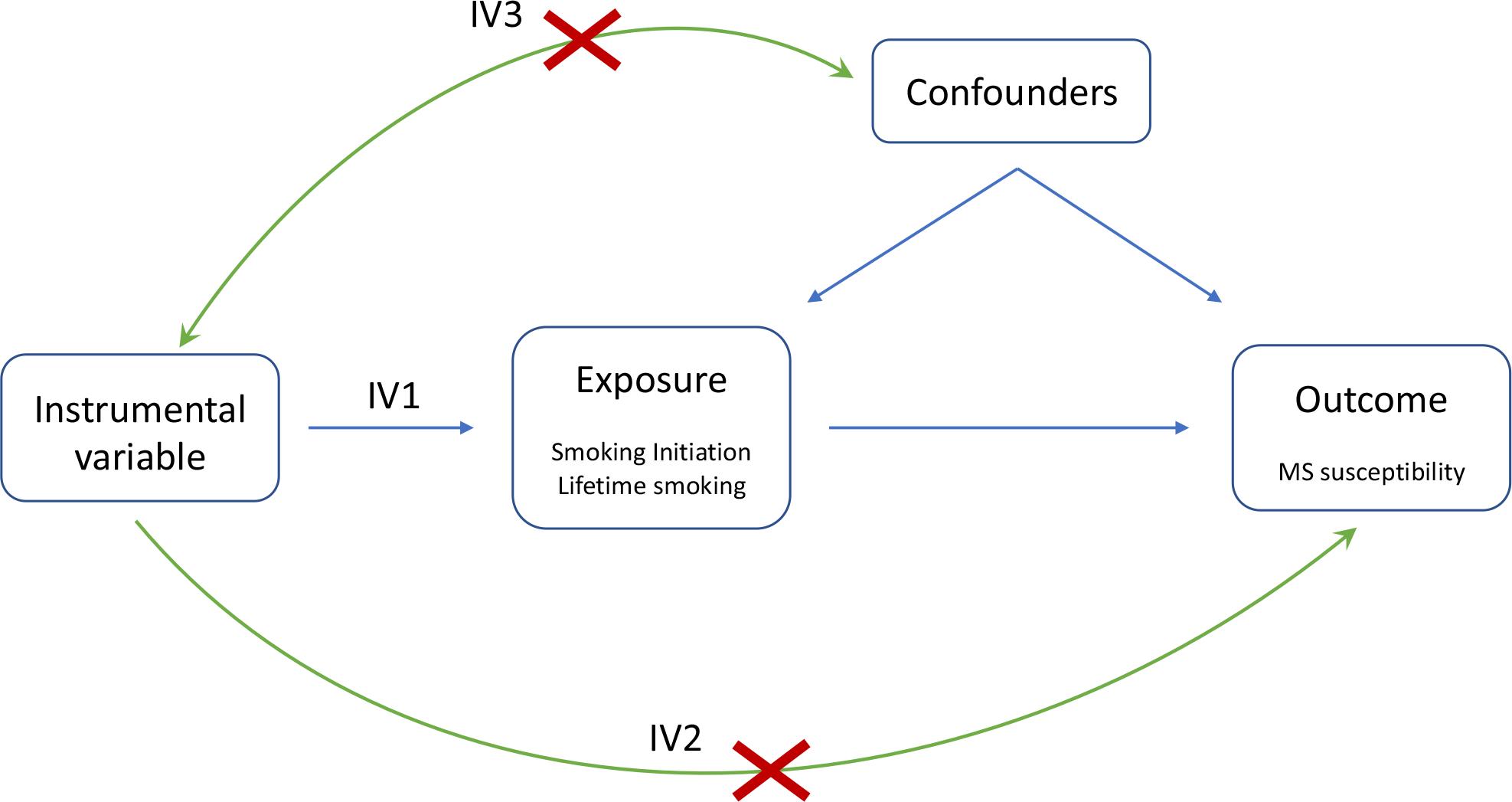
Directed acyclic graph of the Mendelian randomization framework investigating the causal relationship between smoking and multiple sclerosis. Instrumental variable assumptions: IV1: the instruments must be associated with the exposure; IV2: the instruments must influence MS only through smoking; IV3: the instruments must not associate with measured or unmeasured confounders in the smoking to MS relationship.

## Results

### Smoking initiation

The inverse-variance weighted MR estimate (OR 1.03, 95% confidence interval (CI) 0.92 − 1.16) revealed no strong evidence for a causal effect of the genetic risk of smoking initiation on incidence of MS (Fig 3). This was consistent across all MR methods employed, providing further support for the result as each MR method has different assumptions and therefore tests for different violations of those assumptions. Indeed, the weighted median and weighted mode only allow SNPs in the largest homogeneous cluster to contribute to the overall estimate and provide estimates with confidence intervals overlapping the null (Fig 3 and Supplementary Fig 1). The 371 SNPs used as genetic proxies for smoking initiation (Fig 2a and Supplementary Table 1) had an F statistic of 44.90 indicating a strong instrument and that weak instrument bias was unlikely to be influencing the effect estimates. There was evidence of heterogeneity with a large Cochran’s Q statistic of 559.48, p=6.65×10^−6^ and the MR-PRESSO global test value of 562.12, p<0.000125. However, this not indicative of directional horizontal pleiotropy given the consistent MR Egger estimate (OR 1.13, 95% CI 0.67 to 1.91), small intercept (0.0017, p=0.73) and symmetrical funnel plot (Supplementary Fig 2). Similarly, MR-RAPS is robust to systematic and idiosyncratic pleiotropy, accounting for weak instruments, pleiotropy and extreme outliers, and gave a similar causal estimate (OR 1.05, 95% CI 0.93 to 1.17). Furthermore, MR-PRESSO removes individual SNPs that contribute to heterogeneity disproportionately more than expected in order to reduce heterogeneity. The MR-PRESSO outlier corrected causal estimate was 1.040 (95% CI 1.040 to 1.041). Therefore, the second IV assumption (known as the exclusion restriction assumption) of MR has not been violated and directional pleiotropy is unlikely to be biasing the estimates, even though the outlier removal automatically leads to over precise estimates. Leave-one-out and single SNP analyses (Supplementary Fig 3 and 4) were conducted as sensitivity tests sequentially omitting one SNP at a time and performing MR using a single SNP respectively to assess the sensitivity of the results to individual variants. These indicated that there is not a single SNP driving the association whose effect is being masked in the overall analysis. The exclusion of exposure variants located within the MHC did not alter the null association between smoking initiation and incidence of MS (Supplementary Table 2).

**Fig 2.**
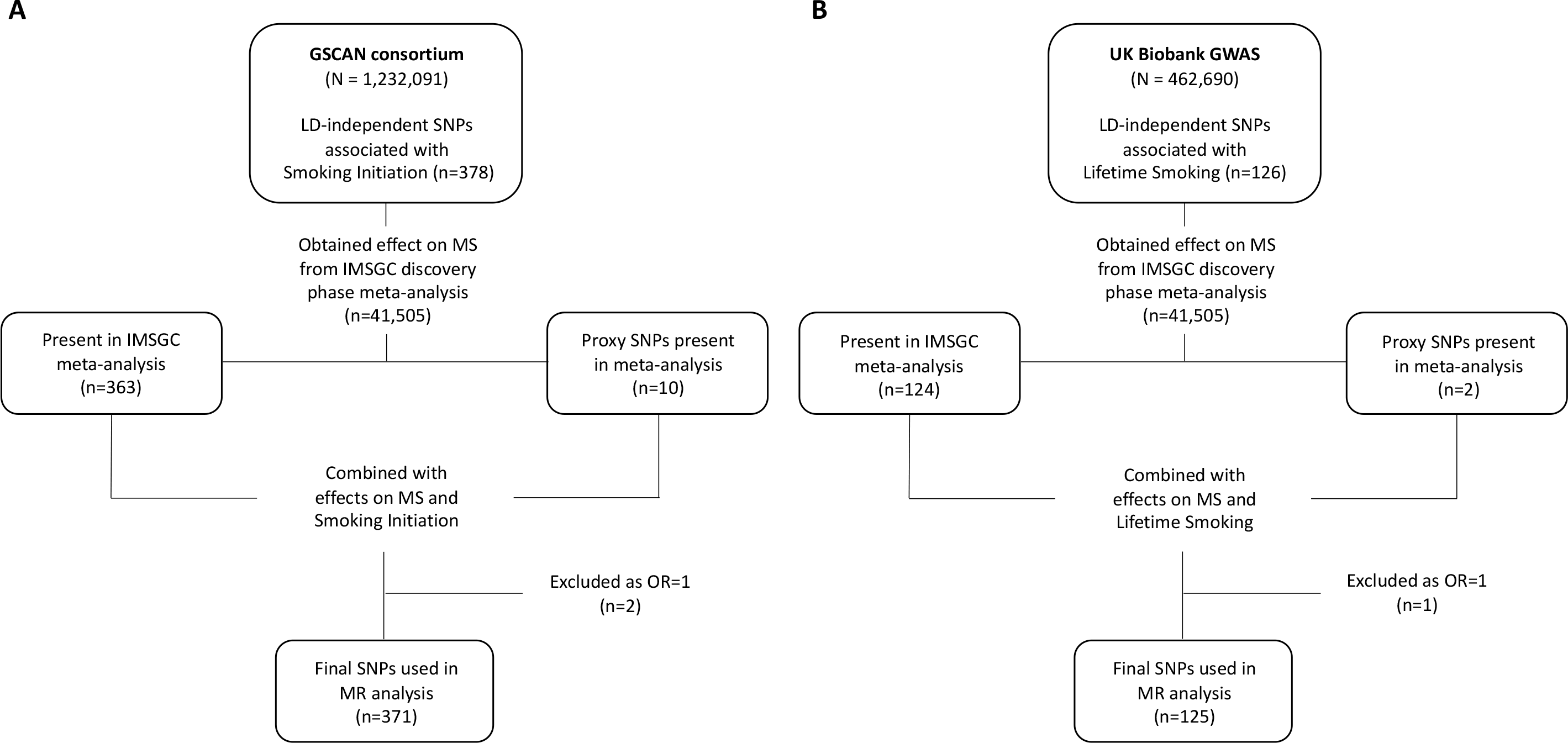
Flowchart for selection of genetic variants associated with smoking initiation (A) and lifetime smoking (B). Abbreviations: GSCAN: GWAS & Sequencing Consortium of Alcohol and Nicotine use; GWAS: Genome-Wide Association Study; LD: Linkage Disequilibrium; MS: multiple sclerosis; IMSGC: International Multiple Sclerosis Genetics Consortium; MR: Mendelian Randomization; SNP: Single-Nucleotide Polymorphism.

**Fig 3:**
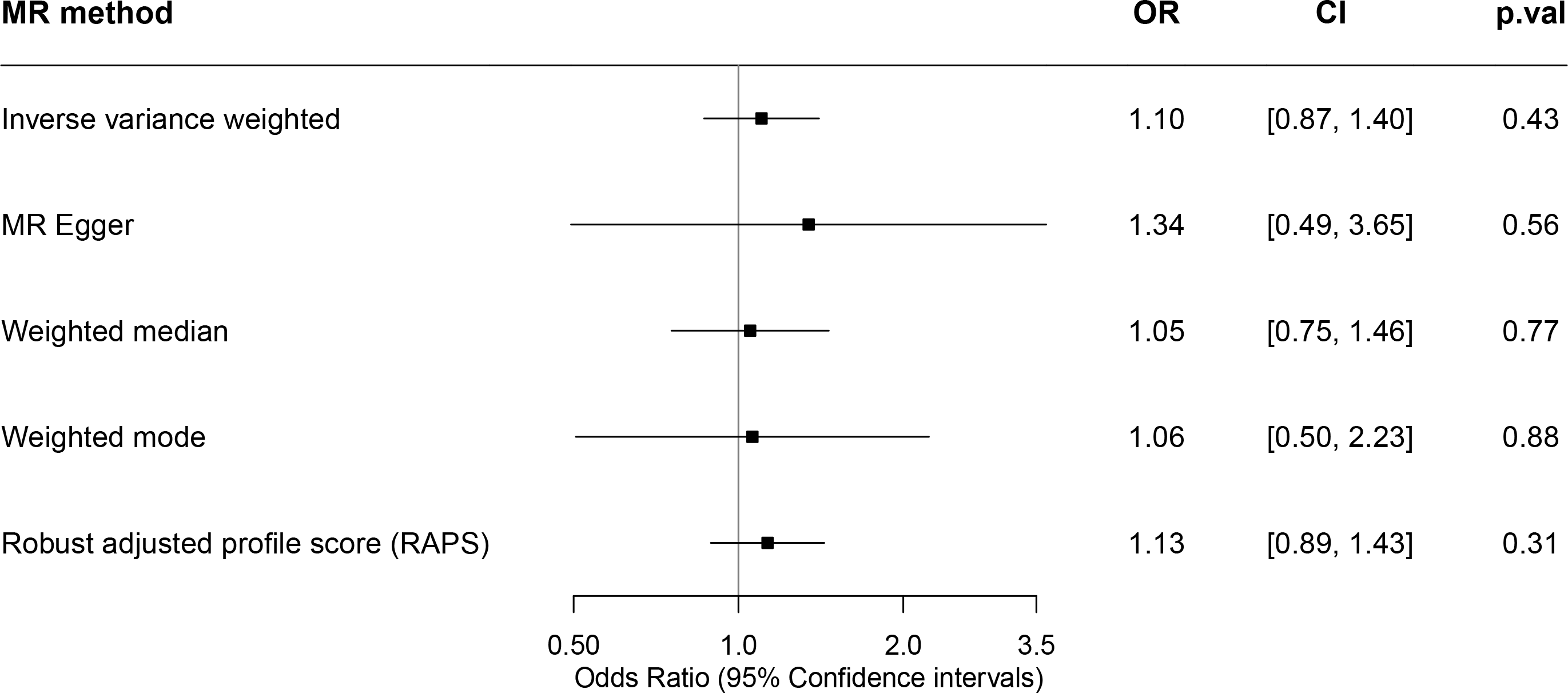
Two-sample Mendelian Randomization estimates of the association between smoking initiation and incidence of multiple sclerosis. Odds ratios are expressed per unit increase in log odds of ever smoking regularly (smoking initiation). MR: Mendelian Randomization; OR=Odds Ratio; CI=Confidence Intervals; p.val: p value.

### Lifetime smoking

There was no clear evidence for a causal effect of the genetic risk of lifetime smoking on incidence of MS (Fig 4). The 125 SNPs used as genetic proxies for lifetime smoking (Fig 2b and Supplementary Table 3) had an F statistic of 44.05 indicating a strong instrument that is unlikely to cause the effect estimates to be affected by weak instrument bias. The inverse-variance weighted MR analysis estimate (OR 1.10, 95% CI 0.87 to 1.40) revealed no strong evidence for a causal effect of the genetic risk of lifetime smoking on incidence of MS and was consistent across all MR methods employed (Fig 4 and Supplementary Fig 5). There was evidence of heterogeneity among the individual SNP effect estimates for lifetime smoking with a large Cochran Q statistic (156.18, p=0.02) and MR-PRESSO global test estimate of 158.4895, p=0.03. However, this was not supported by the symmetrical funnel plot (Supplementary Fig 6) nor by any outliers detected in the MR-PRESSO test. Furthermore, the small MR Egger intercept (−0.003, p=0.69) and consistent MR Egger estimate (OR 1.34, 95% CI 0.49 to 3.65) suggests that the magnitude of potential bias from directional pleiotropy is low. Furthermore, there was no single SNP driving the association whose effect is being masked in the overall estimate as demonstrated by the leave-one-out and single SNP sensitivity analyses (Supplementary Fig 7 and 8). MR excluding the lifetime smoking associated variant located within the MHC region yielded consistent results overlapping the null (Supplementary Table 4).

**Figure 4:**
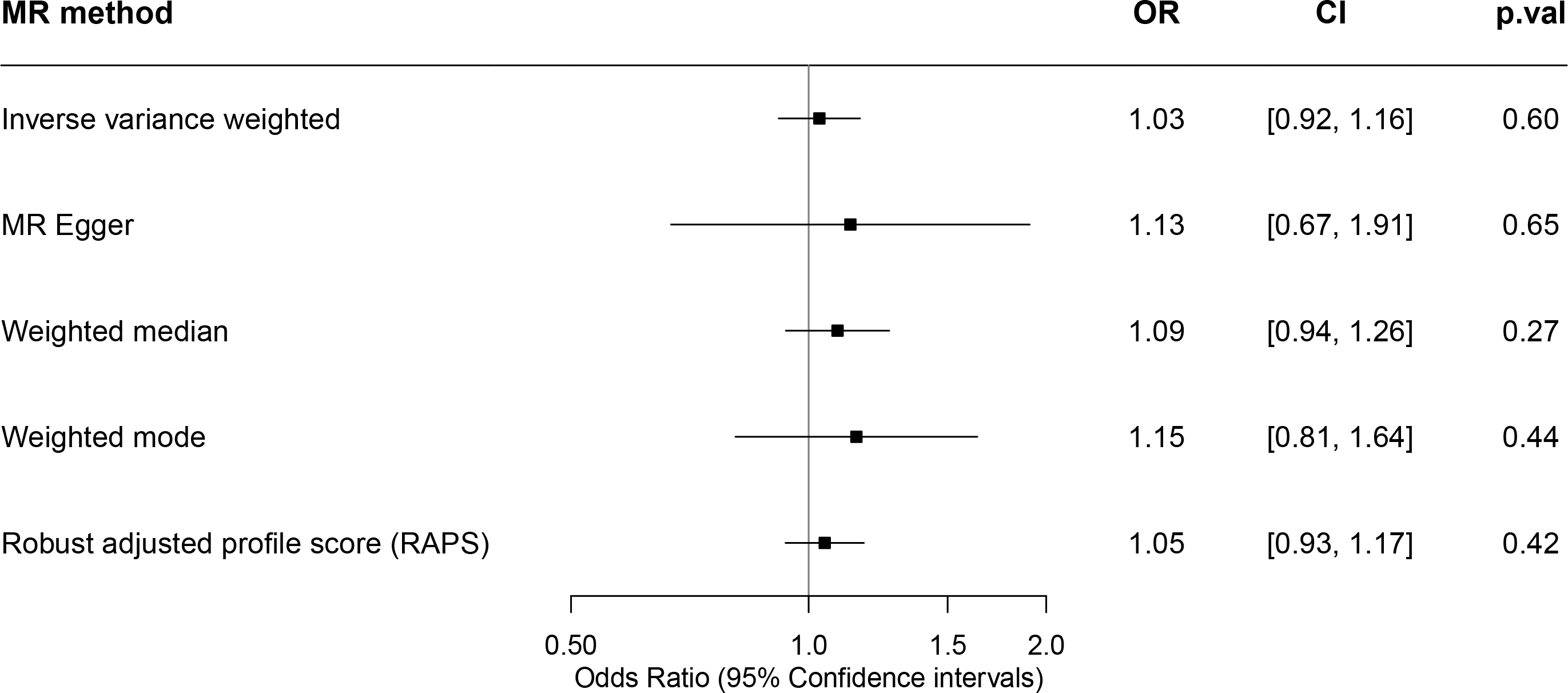
Two-sample Mendelian Randomization estimates of the association between lifetime smoking and incidence of multiple sclerosis. Odds ratios are expressed per 1 standard deviation increase of the lifetime smoking index. MR: Mendelian Randomization; OR=Odds Ratio; CI=Confidence Intervals; p.val: p value.

A bidirectional analysis shows that there was no clear evidence that a genetic predisposition to MS is associated with either smoking initiation or lifetime smoking (Supplementary Table 5 and 7). MR of MS associated variants located within the MHC region yielded consistent results overlapping the null (Supplementary Table 6 and 8).

## Discussion

This study uses the MR method to estimate the causal effect of smoking on risk for MS. Using a two-sample MR design in 14,802 MS cases and 26,703 controls, we found little evidence that both genetically predicted smoking initiation and lifetime smoking are associated with MS risk. These findings suggest that smoking is not a clear environmental risk factor for MS susceptibility and are in line with a recent independent study (27). Although a small effect cannot be entirely excluded, the relatively narrow confidence intervals, particularly for smoking initiation, make a clinically relevant effect less likely.

This contradicts previously reported observational studies that show an association with MS risk among smokers, compared to non-smokers, of a meta-analysed effect estimate odds ratio of 1.5 (4,11). The studies included limitations such as self-report MS diagnosis (28), participation rate less than 80% (29–31) and loss to follow up (32). Additionally, observational studies may have heterogenous results due to how smoking status was defined (11). The strength of association and causality between smoking and MS risk has been suggested due to a dose-dependent relationship in duration and intensity of smoking (4,33) as well as from the interaction between compounds present in cigarettes and specific genetic HLA variants, which include the presence of HLA-DRB1*15, the absence of HLA-A*0201 (34) and specific N-acetyltranferase 1 (NAT1) polymorphisms (35). These genetic variants facilitated epitope cross-reactivity and activation of T cells and smoking may strongly influence the risk of MS observed with these HLA genotypes. However, other studies have failed to replicate this interaction (36,37). In order to test this interaction in an MR casual inference context, a factorial MR design in MS patients with and without those alleles would be required. This was not possible in the present study due to the use of GWAS summary statistics. Observational estimates may have also been biased by residual or unmeasured confounding from factors influencing both smoking status and MS. For example, co-morbidities and socioeconomic status may influence the likelihood of being a smoker and having MS (15,38).

Reverse causation could also partly explain the discrepancy between our MR results and observational studies especially as MS onset may occur long before the first clinical symptoms (39). For instance, this prodromal phase is characterized in part by a higher risk of depression and anxiety up to 10 years prior to MS diagnosis (40), and these in turn are associated with a higher rate of smoking. This study sought to reduce bias from confounding and reverse causation by using a MR design given genetic variants are much less associated with confounders than directly measured environmental exposures (41) (here smoking) and genetic variants are fixed over our lifetime ensuring directionality of effect. This is a major strength of this study in establishing causality in the relationship between smoking and MS risk. Additionally, MR reverse direction MR was performed and shows that reverse causation is unlikely to be playing a role. A further strength of this study is the use of robust genetic instruments which are strong predictors of smoking behaviour. Finally, we used multiple MR methods and sensitivity analyses to test for bias from directional horizontal pleiotropy. Our estimates were consistent across these multiple methods, strengthening our conclusions.

The current study cannot inform us about the effects of smoking on MS symptom severity, disability or progression of disease. Indeed, smoking shows an association with disease progression, disease activity (new lesions on magnetic resonance imaging (MRI); clinical relapse rates) and brain atropy (15). Observational studies have shown an association between smoking and progression from relapsing remitting MS to secondary progressive MS with a dose-response relationship (42–46) as well as a faster rate increasing Expanded Disability Status Scale (EDSS) (22). However, more research in this area is needed to for a definitive conclusion of an effect and specific mechanisms of action. As new methods are being developed to assess disease progression using MR (47), when a GWAS of MS progression becomes available, future studies should explore the association between smoking and the different measures of MS progression in a MR framework.

The instrument predicting smoking initiation and lifetime smoking were broadly distinct (only 9 SNPs overlapping). The measure of lifetime smoking exposure takes into account smoking status and, among ever smokers, duration, heaviness and cessation. Although our lifetime smoking instrument captured smoking heaviness in part, however we were unable to explore whether there was a dose-response relationship between the number of cigarettes smoked and the likelihood of developing MS given we were unable to stratify the MS GWAS by smoking status. Most, but not all (30,48,49), evidence to date seems to suggest that there is a positive correlation between the amount smoked and the severity of illness (4,31,37,43,50–53). It might be that rather than a causal relationship between smoking and MS risk, that smoking instead accelerates the disease process in those that would have already developed MS.

Limitations of this study are, firstly, that although we assessed pleiotropy using MR methods that account for pleiotropic effects, pleiotropy can only be addressed indirectly, and some SNPs may relate to MS risk through pathways other than smoking. We did not find evidence for bias for horizontal pleiotropy using the MR Egger intercept test nor the funnel plots which did not reveal evidence of directional, or unbalanced, pleiotropy. Secondly, this study was a stwo-sample MR using MS meta-analysis summary statistics and therefore this does not allow for gene-environment interaction or sex stratified analysis.

In conclusion, we find no clear evidence for a causal effect of smoking on the risk of developing MS. Previous observational results may have been due to confounding factors, which we have avoided through our analysis. Future research should focus on the effect of smoking on the disease course of MS and its effect on progression.

## Methods

### Genetic instruments for smoking

#### Smoking initiation

We used the most recent GWAS of smoking initiation from the GWAS & Sequencing Consortium of Alcohol and Nicotine use (GSCAN) consortium which identified 378 conditionally independent genome-wide significant SNPs in a sample of 1,232,091 individuals of European ancestry. These genetic variants explain 2% of the variance in smoking initiation(25).

#### Lifetime smoking

In order to incorporate measures of smoking heaviness without having to stratify on smoking status (which is not possible in the two-sample MR context without a stratified GWAS of MS), we used the GWAS of lifetime smoking conducted in 462,690 individuals of European ancestry from the UK Biobank (26). Lifetime smoking is a combination of smoking initiation, duration, heaviness and cessation described in detail elsewhere (26). This GWAS identified 126 independent genome-wide significant SNPs that explain 0.36% of the variance(26).

### Genetic variants associated with multiple sclerosis

Effect estimates and standard errors for smoking associated SNPs on MS susceptibility were obtained from the summary statistics of the discovery cohorts of the latest International Multiple Sclerosis Genetics Consortium (IMSGC) meta-analysis, including 14,802 cases and 26,703 controls (54). All details relating to demographic characteristics, MS case ascertainment and eligibility criteria for the meta-analysis can be found in the original publication (54). For SNPs not available in the IMSGC dataset, we identified proxy SNPs in high linkage disequilibrium (r^2^ > 0.8) using an online tool LDlink [https://ldlink.nci.nih.gov/?tab=ldproxy], giving a total of 371 SNPs for smoking initiation instrument and 125 SNPs for lifetime smoking (Figure 2 and Supplementary Table 1 and 2).

### Mendelian Randomization analyses

A two sample MR was undertaken to obtain effect estimates of genetically predicted smoking on MS susceptibility, using both initiation and lifetime proxy measures. MR and sensitivity analyses were performed in R (version 3.5.1) using the TwoSampleMR R package (https://mrcieu.github.io/TwoSampleMR/) (55) with effect estimates compared across five different methods: inverse variance weighted (IVW); MR Egger (56); weighted median (57); weighted mode (58); robust adjusted profile score (RAPS) (59); pleiotropy residual sum and outlier (PRESSO) (60). Given the different assumptions that each of these methods make about the nature of pleiotropy, consistency in the point estimate across the methods strengthens causal evidence (61). The IWV method is the main analysis and the other methods provide sensitivity analyses. Instrumental variable analysis of MR is based on a ratio of the regressions of the genetic instrument-outcome association (weighted smoking associated SNPs with MS from IMSGC) on the genetic instrument-exposure association (smoking associated SNPs with smoking initiation or lifetime smoking in the independent smoking GWASs). For smoking initiation, the odds ratios are expressed per unit increase in log odds of ever smoking regularly (smoking initiation); for lifetime smoking, the odds ratios are expressed per 1 standard deviation increase of the lifetime smoking index.

Additional sensitivity analyses were performed in order to formally test for potential violations of MR assumptions. The mean F statistic was calculated as an indicator of instrument strength (a value of >10 indicates a strong instrument) and the Cochran’s Q statistic was assessed as a measure of heterogeneity for the IVW method to estimate whether the individual SNP effects of smoking on MS were inconsistent. The MR Egger intercept was assessed to detect directional pleiotropy where the genetic instruments would be influencing MS through another pathway other than smoking. To identify potentially influential SNPs, which could be driven for example by horizontal pleiotropy, we used leave-one-out and single-SNP MR analyses. Additionally, due to the strong genetic signal for MS within the MHC region and high potential for pleiotropy, MR analysis excluding exposure variants located within the extended major histocompatibility (MHC) region was performed (defined as base positions 24,000,000 to 35,000,000 on chromosome 6 [GRCh37].

## Data Availability

The data underlying the results presented in the study are available from the GWAS studies cited.

